# Simulating Retarded SEIRS model for COVID-19: will the second epidemic happen?

**DOI:** 10.1101/2020.12.24.20248842

**Authors:** Hamid Pour Mohammad

## Abstract

In this paper, we want to simulate the COVID-19 epidemic according to the Retarded SEIRS model. One of the main questions in the human mind is whether the COVID-19 epidemic will happen again. Therefore, a criterion must be set for the occurrence or non-occurrence of the disease. With the Retarded SEIRS model and this criterion, we can predict whether the Covid-19 will re-emerge. So far, a large number of researches that have been presented in scientific groups or communities have been based on the SIR or SEIR model. But we assume that each recovered individual is immune to the disease for a limited time, and then will be susceptible again. As we know, this assumption was also true for SARS.

## 1. Introduction

From time immemorial until now, various diseases have threatened the human race, the most influential of which in recent decades is COVID-19. The effects of this disease have been extended to economic and political fields. Since COVID-19 can spread significantly in the world, we focus on studying its recurrence after the first pandemic. Various models have been studied for this disease, the most important of which are *SEIR* and *SIR* But in fact, the question that arises here is whether it is possible for the COVID-19 to become re-epidemic. The answer to this question certainly depends on the model that COVID-19 follows.

In general, if a person is patient with a disease, three conditions may occur after recovery:

1. Immediately after recovery, he/she is prone to become patient;
2. He/she is immune to the disease for a limited time after recovery, and after that time he/she will be prone to be patient again;
3. Once he/she recovers, he/she will have lifelong immunity to the disease and will never be paatient.

For example, a child can get the tetanus vaccine and be immune to the disease for a long time, but over the years, that immunity wears off and he/she has to get the vaccine again. Many adult immunization schedules also suggest that the tetanus and diphtheria vaccine, called *Td vaccine*, be given every 10 years. It should be noted that research shows this period of time can be significantly corrected [1]. The problem is whether such a question can also exist for the immunity of the COVID-19.

The details of this disease are still unknown and the related questions can hardly be answered. On the other hand, it is not logical for human beings to wait and find the answer to this question in the future, based on experience. Because today, possible events can be obtained through various simulations. We want to obtain the condition for the recurrence of COVID-19 epidemic through computer simulation. The model we want to use is called *Retarded SEIRS*, and we believe that this model is more realistic than conventional *SEIRS* models. Here, after studing the parameters and simulating the model, we calculate the minimum value required for the temporary immunity duration of COVID-19, to prevent the occurrence of the second epidemic.

## 2. Retarded SEIRS model description

When a person becomes ill with COVID-19 disease, he/she will either die or recover. And if he/she recovers, one of the three above conditions will occur. So far, studies on conditions 1 and 3 have been performed. Therefore, we put our focus on the second one. To examine this condition, we assume that the recovered individuals lose their immunity over a period of time *T* and become fully susceptible to disease. One of the most important questions is, if we make such a presumption for the spread of the disease, then how far are we from the re-emergence of COVID-19?

To model this phenomenon, We first divide people into four categories: susceptible, exposed, infected, and recovered. Because a significant number of researches (such as *Yang et al*. (2020) and *Lopez and Rodo* (2020)) that have been proposed for this disease, have been based on *SEI R* model. However, in this study, it should be considered that the recovered individuals return to the susceptible category (*S*) after the time duration *T* Therefore, according to the sequence of different categories, this model can be called *SEIRS* for short. But since we have not used conventional methods to transfer individuals from recovered (*R*) to susceptible (*S*), we have called it *Retarded SEIRS* model. Because we are exactly using the time latency, not transfer rate-based methods. To examine the changes of categories in the population over time, the dynamic equation for each category must be written. It is clear that the change in the proportion of each category can happen in two ways: either some individuals in a category leave the category and go to the next category, or some individuals of a category die or born. With these interpretations, we have assumed the dynamic equations for categories as equations 1 to 4:

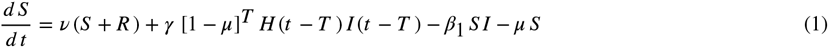

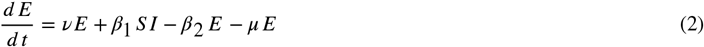

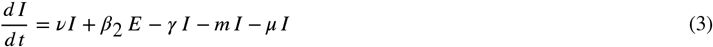

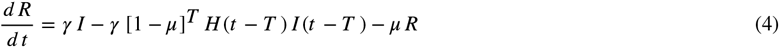

In equation 1, the term related to the births of *S* the *R* and categories is shown as *ν* (*S* + *R*) *ν* so that is the birth rate; in other words, even if a person is born in *R* category, he/she will be considered susceptible to the disease. The second term in equation 1 (ie, *γ* [1 − *μ*]^*T*^ *I* (*t* − *T*) *H* (*t* − *T*)) refers to recovered individuals who have returned to the category of susceptible individuals after the duration *T* has elapsed since their recovery. In other words, this term is subtracted from equation 4 and added to equation 1. The explanation of this term is given in detail in the description of equation 4. Parameter *T* will be very important in this model, as it expresses the duration of temporary immunity of each individual after getting recovered. The third sentence is −*β*_1_ *SI*, which indicates the transmission of individuals from to with the rate *β*_1_, due to exposure of individuals to *I* Also, in the fourth term, individuals will die at the rate *μ*.

In equation 2, it can be seen that the birth of infans in the category *E* will occur at the rate and as the form *vE*. The second term in this equation states *S* individuals that have been exposed to *I*, so they will be added to *E* category as *β*_1_ *S I*. Then in term −*β*_2_ *E*, these individuals enter the infected category at *β*_2_ rate from *E* category. To be more precise, this term is related to the departure of *E* individuals from the latent period of the disease and their entry into the infected category. The last term, like other categories, indicates the natural death of individuals.

Now it will not be difficult to interpret equation 3. Term *vI* states that the birth of each infant in the infected category means that the infant is infected too. We have not seen this to be true yet, although it seems very reasonable (similarly, we assumed that the share of births in category *E* would be added to *E*). The description of the second term is also given in the paragraph above; but the third term (− *γ I*) is about infected recovering at *γ* rate and entering *R* category. Although we emphasize that a number of infected individuals will lose their lives; these individuals, whose share is included in the fourth term (ie − *ml*), die at rate and simply because of COVID-19.

Equation 4 is a very basic equation in this model. Till now, almost all transfers between categories have been controlled by rates (except for the second term in the first equation, which we are going to explain here). In the first term (*γ I*) we see that infected individuals have entered *R* category at γ rate. But the second term is written in a way independent of the rates. Being independent of the transition rate means that the rate 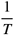 will not be used to transfer individuals from *R* to *S* As mentioned, the second term in equation 4—ie, − *γ* [1 − *μ*]^*T*^ *I* (*t*− *T*) *H* (*t* − *T*)—refers to recovered individuals who, after passing the duration *T* from the moment of their recovery, have returned to the category of susceptible individuals. These are the infected individuals who were excluded from *I* category at time *t* − *T* and entered R category at the rate *γ*. Also, the presence of Heaviside function H guarantees the transition after the duration *T*. For this reason, the share of these individuals at the moment *t* is subtracted from equation 4 and added to equation 1. The term [1 − *μ*]^*T*^ may sound a little unfamiliar, but note that some of these recovered individuals will die during the duration *T*, due to the natural death rate *μ* per unit time. Therefore, only [1 − *μ*]^*T*^ fraction of the initial number of these individuals will be alive. We must note that 1 and *μ* have the same dimension. In this model, there is an important difference compared to conventional *SEIRS* models; in conventional models, there is a rate for transferring individuals from *R* to *S*, which is basically the same as 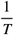. Recovered will then enter the category of the susceptible with the term 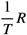. But we believe that this will have unrealistic consequence. Note that *T* is much larger than the other time durations of COVID-19 in these equations—such as latent period, recovery period, etc. We also know that after the outbreak or even during it, a significant portion of the population falls into the R category. In fact, it is clear that as R goes, so doeS 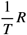 term, and so a flood of recovered individuals will flow to the susceptible category. After R reaches its maximum value, this flood also reaches its maximum and then will decrease. But This event is completely unrealistic. Because we expect that even if a significant number of infected enter R category, they have to wait for their temporary immunity T to expire, and then enter S category. however, in conventional *SEIRS* models, as soon as these individuals enter R, the flow of them to the S will be at its maximum. So we explicitly suggest using − *γ* [1 − *μ*]^*T*^ *I* (*t* − *T*) *H* (*t* − *T*) term instead of the 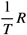. Such corrections can be applied to the other terms—although we did not, but it is better to correct them to make the model more realistic. We also note that the nature of “temporary immunity” is well considered in this method. But this was not the case with conventional *SEIRS* models.

## 3. Calculating the parameters

We now describe the parameters in equations 1 to 4. To obtain the natural birth and death rates, we used a research conducted by the United Nations [4]. This study shows that the average number of alive births per woman (during life) is 2.5 children. Since life expectancy at birth of women in 2019, is reported to be 75.0 years [4], we estimate our required birth rate as:

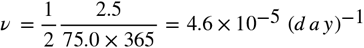

Note that we ultimately intend to investigate the normalized population, and since women make up almost half of the world population, the coefficient 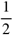 would be justified. On the other hand, life expectancy at birth—for both sexes, in 2019—is 72.6 years. This value causes us to estimate natural death rate:

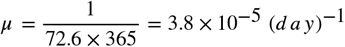

for whatever reason except COVID-19.

According t o *Li et al*. (2020), the latent duration lasts 5.2 *days* (95 % *C I*, 4.1 *t o* 7.0) days, which results in *β*_2_ = 0.19 (*day*)^−1^ (95 % *CI*, 0.14 − 0.25) infection rate. The recovery time of each infected individual is 24.7 (*days*) (95 % *Cr I* 22.9 − 28.1) [6] and therefore the recovery rate will be *γ* = 0.040 (*d a y*)^−1^ (95 % *C r I* 0.035 − 0.044). The share of deaths due to COVID-19 is equal to .38 % (95 % *Cr I* 1.23 − 1.53) [6], which causes the fatality rate to be estimated to be:

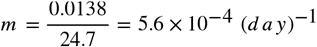

On the other hand, the basic reproductive number of this disease is estimated to be *R*_0_ = 2.68 (95 % *C r I* 2.47 − 2.86) [7]. Given that *R*_0_ in this model is obtained from the definition in equation 5 [8], the transmission rate *β*_1_ can be estimated at 0.11 (*day*)^−1^ using the above results. Note that we exactly used the definition of *R*_0_ in *SEIR* model, which would be a logical calculation.

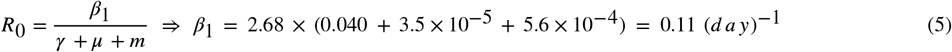

## 4. Simulating the dynamics of disease outbreak

The equations 1 to 4 can be solved by different numerical methods, and we chose Fourth Order Runge-Kutta for this purpose. Before solving these equations, we emphasize that we have first normalized the population to 1. It means 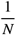 of population is infected and the rest—which is 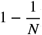) —are susceptible. *N* is the total number of people in the world. In this text, we consider the initial proportion of infected to be:

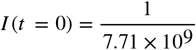

which is equivalent to the proportion of one person among the total world population. Because the world’s population in 2019 was *N* = 7.71 × 10^9^ [9]. Also, at the beginning of the outbreak, we consider the proportions of *E* and *R* to be zero. On the other hand, due to births and deaths, the initially normalized population will not remain 1. So we have to keep in mind that if, for example, after a long time, the individuals in category *S* have a proportion of 0.4, it means that the ratio of their population to the initial population of the world (in 2019) is 0.4. So when we talk about *S, E, I* and *R* proportions, we mean the ratio of the number of individuals in each of these categories to the total initial population. In other words:

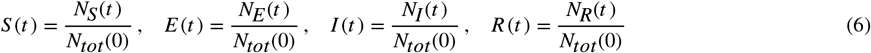

Our intention is to calculate the number of infected individuals for different amounts of *T* As the point is clear, our method is numerical and deterministic. It should be noted that *S, E, I* and *R* values can not be negative. It should also be noted that the number of individuals in a category cannot be reduced to any desired positive number. For example, after the first outbreak subsides, the proportion of *I* individuals cannot be reduced to 10^−12^. Because none of the human societies—even the largest of them—have a population of 10^12^, and so we can’t be in a situation where relatively one individual in 10^12^ people is infected. In other words, given the world’s population, which is the largest imaginable society, the proportion of each category must be such that the number of its individuals can be considered comparable to the lowest positive population proportion of humans in the world—which corresponds to one person in the world. Otherwise, we consider the proportion of that category to be zero. we will provide a more comprehensive explanation in this regard.

We see the result of one of COVID-19 simulations over fifteen years in figure 2. As can be seen from figures (2. *A*) and (2.*B*), with the outbreak of the disease, the significant portion of the population becomes infected and then the proportion of infected (*I*) decreases. Note that these two figures are related to one simulation and are drawn in two parts just for better observation. In this simulation, it is assumed that the immunity of each recovered individual disappears after 3.0 years. After a significant portion of the recovered individuals have lost their immunity, to which they are added to category, world will be ready for a re-epidemic—ie, the second epidemic. But this re-epidemic will not be so evident until the sixth year. Because after the first epidemic, the number of individuals decreased so much that for several years it could not make a significant fraction of the population infected. Note that this is quite natural given the dynamic equations of the disease and the numerical method used. For example, between the first and second peaks (figure2.*B*), *E* and *I* proportions have the values less than 10^−12^. On the other hand, given the world’s population, we can never face such a proportion of the world’s population, and therefore the disease is practically gone. As a result, the second, third, and subsequent epidemics, which occur at approximately five-year intervals (figure 2), will not occur in the real world.

**Figure 1).**
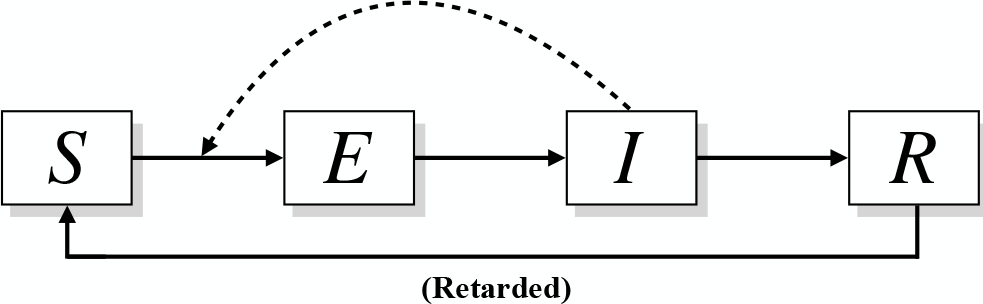
*Retarded SEIRS* model overview. Susceptible individuals will be exposed by infected individuals. Exposed individuals will then enter the infected category after a short time. The infected individuals will recover, and then join the susceptible category again. It should be noted that for better understanding, births and deaths are not included in this figure. But it is clear that the births should be included in some categories; in addition, given that natural death occurs in all categories, another output must be considered for each category. Also, some individuals will die in the infected category due to the fatality of the disease.

**Figure 2).**
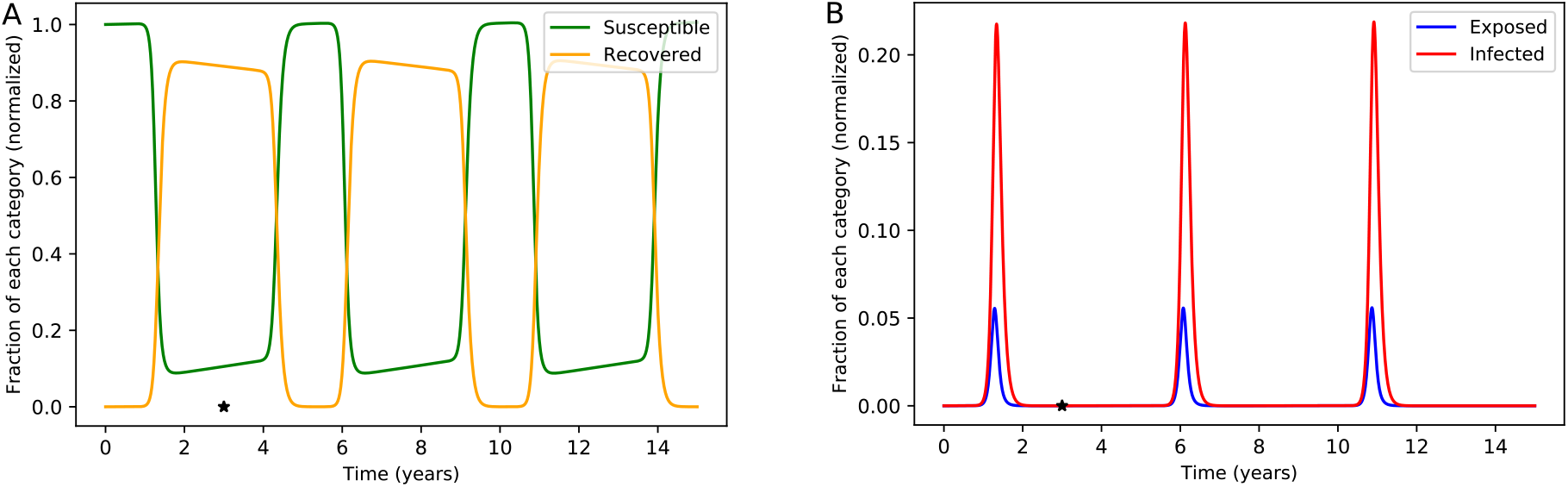
*Retarded SEIRS* Model for COVID-19 epidemics. A) *S* and *R* proportions. B) *E* and *I* proportions. Both figures are for one simulation, and are drawn in two parts because of better representation. Here it is assumed that each recovered individual is immune for *T* = 3.0 *years*. So at *t* = 3.0 *years* and after that, the immunity of individuals who got infected early in the outbreak will be lost. The asterisk shows the first individual who loses the immunity, which corresponds the first infected individual. After the first peak of the epidemic, the proportion of *E* and *I* will decrease sharply and then increase again. Epidemics occur every few years, approximately every five years. It should be noted that each of *E* and *I* proportions sometimes fall below 10^−12^—for example, between the first and second epidemic peaks—and these events cannot be observed in human societies. This simulation shows that the use of numerical methods, regardless of the *S E,I* and *R* values, can lead to unrealistic results. The parameters used in this simulation are are shown in table 1. To view the code related to this simulation, refer to *SEIRS Figure*.2.*py* code (supplemental files).

Another point to consider in the simulations of this text is the calculation of *I* (*t* – *T*) substeps in Fourth Order Runge-Kutta method. As we know, to perform the calculations of each time step, ie from *t* to + Δ*t*, for each of the proportions *S* (*t*), *E* (*t*), *I* (*t*) and *R* (*t*), we must calculate four substeps. But the main problem is that there is no separate equation for *I* (*t* – *T*), which is time-dependent. Hence, we use the calculations we did for *I* at time *t*−*T* In fact, using the substeps we obtained in the past for *I* we affect the substeps of *I* (*t* – *T*). For more details, refer to the code of each simulation for a better understanding. Using this method, delay-dependent calculations of *I* (*t* – *T*) can be performed.

**Table 1).**
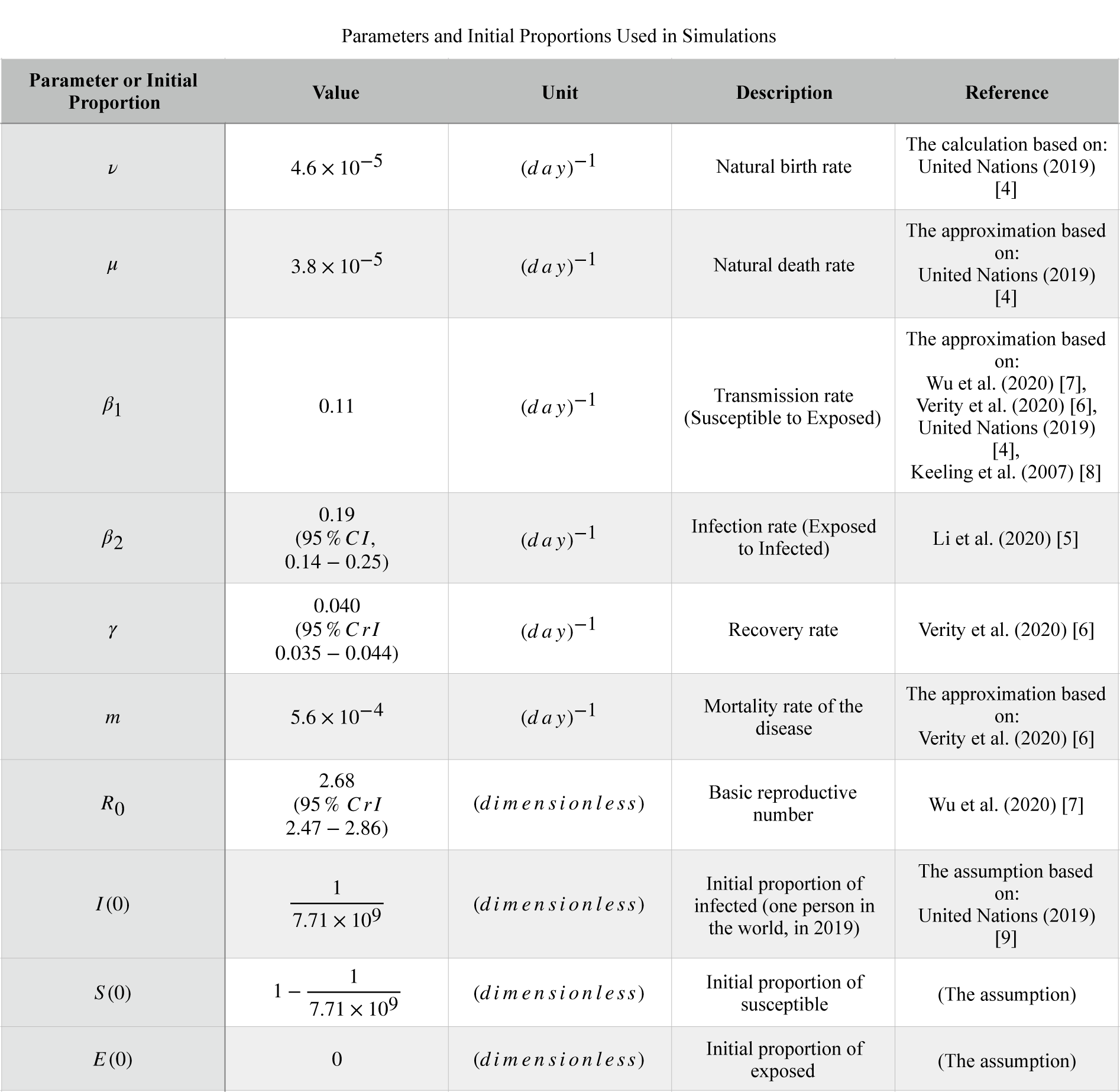
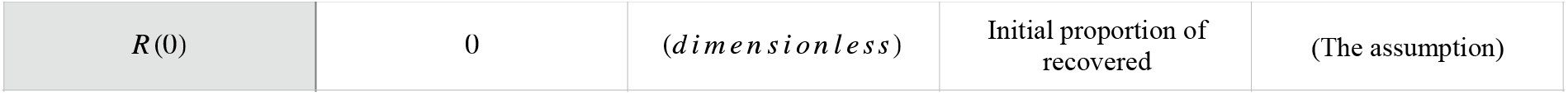
Parameters and initial values used in *Retarded SEIRS* model simulations. Some intervals are shown for some parameters, but we did not use the intervals in our simulations. We only considered the main value of each parameter.

We now turn to the question of what happens if we reduce the duration of temporary immunity (*T*). In this case, *R* individuals stay shorter in the temporary immunity duration, and therefore can get to the *S* category sooner. So we can expect a re-epidemic by adopting a small number for *T*. We show this simulation for *T* = 1.0 *years* in figure 3. The minimum of *I* between the first and second epidemic is 4.27 × 10^−5^, which would be quite significant. Because such a proportion of the world’s population is equal to:

**Figure 3).**
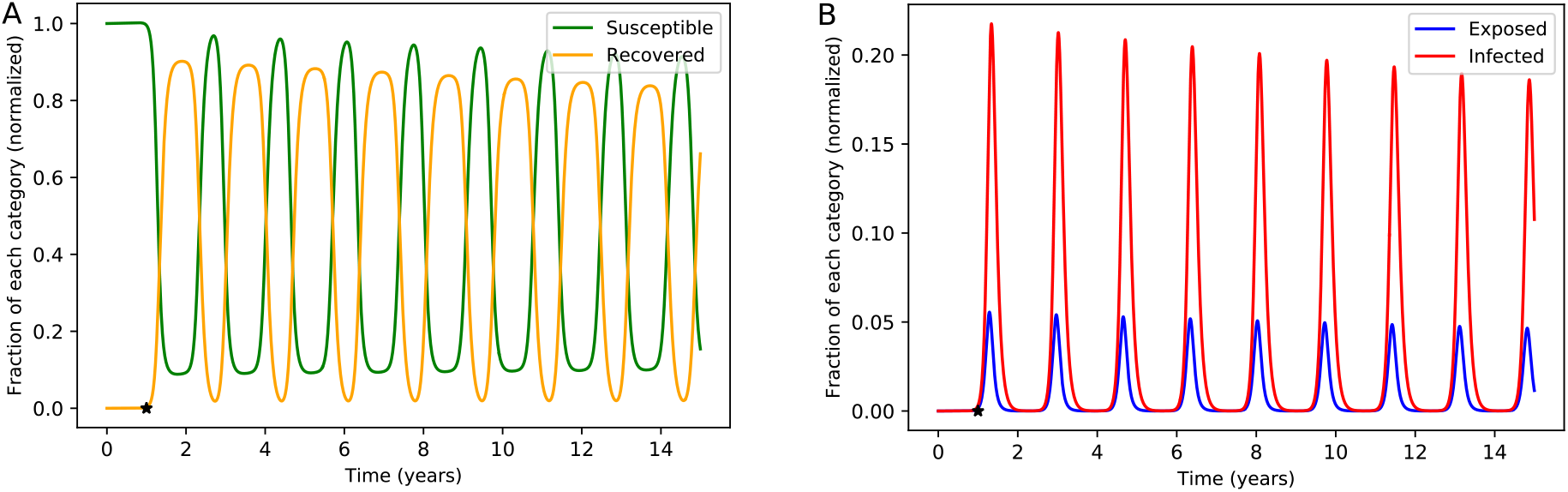
Simulation of *Retarded SEIRS* model, for *T* = 1.0 *years* over fifteen years. A) *S* and *R* proportions. B) *E* and *I* proportions. It is emphasized again that both diagrams are related to one simulation and are calculated for one-year temporary immunity duration. The other parameters are the same as the parameters in table 1. The first, second, and third epidemics (and even after them) can be clearly seen from the curve of *I* (or other categories). To view the code related to this simulation, refer to *Ret arded*. *SEIRS*. *Figure*.3.*p y* code (supplemental files).

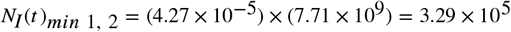

infected individuals. That is, under such circumstances, in the smallest amount, we still face several hundred thousand infected individuals. Then we approach the peak of the second epidemic and then the next stages.

## 5. Finding the threshold of temporary immunity *T* for non-occurrence of re-epidemic

We performed the first simulation with *T* = 3.0 *years* and said that this simulation would not lead to a second epidemic (figure). Then we performed the second simulation with *T* = 1.0 *year* and found that the second epidemic occurs (figure). According to the argument made earlier, we should expect that in a particular *T* there is a threshold for non-occurrence of the second epidemic. This *T* will definitely be between 1.0 and 3.0 years. But the question is, what is our criterion for diagnosing the occurrence or non-occurrence of the second epidemic? Our criterion is defined as follows: if the minimum of between the first and second epidemics—in the simulation—is less than 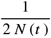, then the second epidemic will not actually occur. *N* (*t*) is the population of the world at time *t* We have run our simulations with an initially normalized population. Also, we know that the smallest imaginable proportion of the population in the real world is 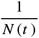 In other words, the largest conceivable society is the entire population of the world, and so if we want to use the smallest conceivable fraction in the real world, we must consider a minimum number for *I* fraction to shrink. The term 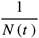 means the share of one person in the total population. If in the simulations the proportion of a category reaches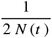, it means that the share of that category—from the point of view of the real world—is equal to half of a human. That is, in the largest imaginable human population, comprising the entire human population of the planet, we are faced with the share of half of a human. It is clear that if we choose a smaller population, we will face a smaller share of humans. On the other hand, in this example, we will be dealing with proportions similar to 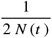 in earlier steps, according to the method we use—Fourth Order Runge-Kutta. If we want to use the probabilistic interpretation for this event, we can say that in these steps we are faced with the product of consecutive probabilities of about 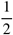. It is clear that if we multiply the numbers close to 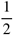 in succession, their final answer will be very small and therefore the probability of that event will be insignificant.

Although our view on this issue is deterministic, it should be noted that when the number of infected individuals is very small, the stochastic effects should also be considered important. These effects may go extinct the disease sooner or later compared to the deterministic perspective. Also, we assumed that the whole world operates symmetrically, and this assumption can be significantly modified.

Now we can look for a particular *T* in the range of *T* = 1.0 *year* to *T* = 3.0 *years*, so that the minimum of *I* between the first and second pandemics in the figure is less than 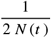 for the first time. By performing simulations for different *T* values in this interval, the threshold of *T* can be easily found. If we do this, we get *T*_*thr*_ = 2.4± 0.1 *years* for the threshold of temporary immunity duration for non-occurrence of re-epidemic. To view the code related to this simulation, refer to *Critical T. p y* (supplemental files). Thus, under this model, COVID-19 pandemic can occur for the second time if its duration of temporary immunity is less than *T*_*thr*_ = 2.4 ± 0.1 *years* In other words, if *T*_*COVID*−19_ < *T*_*thr*_, then the second epidemic will occur. Otherwise, COVID-19 will become extinct, although we have ignored the influence of other factors such as mutations. It should be noted that the current error reported for *T*_*thr*_ is due to the fact that each time we simulated, we changed the value of *T* by 0.1 *year* and then ran the program again. Therefore, the overall error for will be greater than 0.1 *year*, because each parameter in this model also have an error interval in reality, while the intervals are not affected in the simulation.

Now if we focus on the simulation we did for *T* = 2.4 *years*, we conclude that the minimum *I* of between the first and second pandemics is approximately 4.0993 × 10^−11^. We also find that in the previous one and two time steps, we were dealing with approximately 4.0994 × 10^−11^ and 4.1025 × 10^−11^. Such a result was expected. As mentioned earlier, given the numerical method we used, we should expect the numbers obtained for each category in successive time steps to be slightly different. Also, 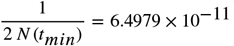 is obtained for the minimum moment; it is clear that all three of the above numbers are less than 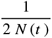.

## 6. An effective suggestion for researchers

We have calculated the threshold of temporary immunity of COVID-19 for non-occurrence of re-epidemic around *T*_*thr*_ = 2.4 *years*. Now we have to wait for the testers to obtain the amount of temporary immunity *T*_*COVID*−19_ experimentally; if *T*_*COVID*−19_ < *T*_*thr*_, we are probably facing a catastrophe. For further investigation, we examined a disease that behaved similarly to COVID-19. A study of 176 patients with Acute Respiratory Syndrome (SARS) found that SARS antibodies persisted for an average of two years. Then in the third year, significant changes occurred in their bodies. Therefore, SARS patient individuals may be re-susceptible to SARS after ≥ 3 *years* [10]. On the other hand, it may be necessary to change the model to individual-based or metapopulation models to achieve more accuracy. Under any circumstances, our suggestion to the medical community is to monitor the first registered patients, because they may become infected for the second time. The sooner we know about the temporary immunity of COVID-19, the better we can react to the recurrence of the disease. We hope to overcome this dilemma with the best efforts of all the people in the world.

## Supporting information

Retarded.SEIRS.Figure.2.py code

Retarded.SEIRS.Figure.3.py code

Critical.T.py code

## Data Availability

All data has been reported according to the mentioned references. Also, the simulation codes are available on GitHub.

https://github.com/chix4030/Simulating-Retarded-SEIRS-model-for-COVID-19-will-the-second-epidemic-happen-

