## Supplementary material for "Simulating Retarded SEIRS model for COVID-19: will the second epidemic happen?": Retarded.SEIRS.Figure.3.py code

December 22, 2020

[ ]:
```
# Retarded.SEIRS.Figure.3.py:
from numpy import zeros
import matplotlib.pyplot as plt

tot=round(365*15) # total time (tot has to be more than 2*T)
T=round(365*1) # temporary immuniny (main)
t=zeros(tot) # time
dt=1 # one day
dt2=dt/2 # half of a day: it's for RK4

# Fractions:
S=zeros(tot)
E=zeros(tot)
I=zeros(tot)
R=zeros(tot)
TT=[i for i in range(tot)]

I[0]=10**-9 *(1/7.71) # initial infected: one person, in the world (in 2019)
S[0]=1-I[0] # initial susceptible

b1= 0.11  # beta_1 (per day) => S to E
b2= 0.19  # beta_2 (per day) => E ti I
v= 4.6 * 10**-5 #nu => per day
u= 3.8 * 10**-5 #mu => per day
uu=(1-u)**T
g=0.040 # gamma => per day
m=5.6*(10**-4) #m => per day
a= 1 # alpha (It is not important yet!)

def H(t): # heaviside function
    if t>=0:
        return(1)
    else: return(0)

iret=[zeros(4) for i in range(tot)] # I_retarded: it's for I(t-T) (because we␣
↪are using RK4 method)
```

```python
# Dynamics:
for i in range(tot-1): # i is equivalent to time => t

    ks1=dt*(v*(S[i]+R[i])+a*g*uu*H(i-T)*I[i-T]-b1*S[i]*I[i]-u*S[i])
    ke1=dt*(v*E[i]+b1*S[i]*I[i]-b2*E[i]-u*E[i])
    ki1=dt*(v*I[i]+b2*E[i]-g*I[i]-m*I[i]-u*I[i])
    kr1=dt*(g*I[i]-a*g*uu*H(i-T)*I[i-T]-u*R[i])

    ks2=dt*(v*(S[i]+ks1/2+R[i]+kr1/2)+a*g*uu*H(i-T+dt2)*(I[i-T]+iret[i-T][0]/2)-b1*(S[i]+ks1/2)*(I[i]+ki1/2)-u*(S[i]+ks1/2))
    ke2=dt*(v*(E[i]+ke1/2)+b1*(S[i]+ks1/2)*(I[i]+ki1/2)-b2*(E[i]+ke1/2)-u*(E[i]+ke1/2))
    ki2=dt*(v*(I[i]+ki1/2)+b2*(E[i]+ke1/2)-g*(I[i]+ki1/2)-m*(I[i]+ki1/2)-u*(I[i]+ki1/2))
    kr2=dt*(g*(I[i]+ki1/2)-a*g*uu*H(i-T+dt2)*(I[i-T]+iret[i-T][0]/2)-u*(R[i]+kr1/2))

    ks3=dt*(v*(S[i]+ks2/2+R[i]+kr2/2)+a*g*uu*H(i-T+dt2)*(I[i-T]+iret[i-T][1]/2)-b1*(S[i]+ks2/2)*(I[i]+ki2/2)-u*(S[i]+ks2/2))
    ke3=dt*(v*(E[i]+ke2/2)+b1*(S[i]+ks2/2)*(I[i]+ki2/2)-b2*(E[i]+ke2/2)-u*(E[i]+ke2/2))
    ki3=dt*(v*(I[i]+ki2/2)+b2*(E[i]+ke2/2)-g*(I[i]+ki2/2)-m*(I[i]+ki2/2)-u*(I[i]+ki2/2))
    kr3=dt*(g*(I[i]+ki2/2)-a*g*uu*H(i-T+dt2)*(I[i-T]+iret[i-T][1]/2)-u*(R[i]+kr2/2))

    ks4=dt*(v*(S[i]+ks3+R[i]+kr3)+a*g*uu*H(i-T+dt)*(I[i-T]+iret[i-T][2]/2)-b1*(S[i]+ks3)*(I[i]+ki3)-u*(S[i]+ks3))
    ke4=dt*(v*(E[i]+ke3)+b1*(S[i]+ks3)*(I[i]+ki3)-b2*(E[i]+ke3)-u*(E[i]+ke3))
    ki4=dt*(v*(I[i]+ki3)+b2*(E[i]+ke3)-g*(I[i]+ki3)-m*(I[i]+ki3)-u*(I[i]+ki3))
    kr4=dt*(g*(I[i]+ki3)-a*g*uu*H(i-T+dt)*(I[i-T]+iret[i-T][2]/2)-u*(R[i]+kr3))

    S[i+1]=S[i]+(1/6)*(ks1+2*ks2+2*ks3+ks4)
    E[i+1]=E[i]+(1/6)*(ke1+2*ke2+2*ke3+ke4)
    I[i+1]=I[i]+(1/6)*(ki1+2*ki2+2*ki3+ki4)
    R[i+1]=R[i]+(1/6)*(kr1+2*kr2+2*kr3+kr4)

    iret[i][0]=ki1
    iret[i][1]=ki2
    iret[i][2]=ki3
    iret[i][3]=ki4 # (not important)


TT=list(map(lambda t:t/365, TT))
```

```
print('Normalizad, RK4:')
print(f'temporary immuniny = {round(T/365,2)} (years)')
print('I_0=(1/7.71) * 10**-9')
print(f'b1= {b1}  # beta_1 (per day) => S to E')
print(f'b2= {b2}  # beta_2 (per day) => E ti I')
print(f'gamma= {g} (per day)')
print(f'mortality rate= {round(m*10**4,2)} * 10**-4 (per day)')
print(f'v= {v*10**5} * 10**-5 #nu => per day')
print(f'u= {round(u*10**5,1)} * 10**-5 #mu => per day')

# S & R:
plt.plot(TT,S,label='Susceptible', color='green')
plt.plot(TT,R,label='Recovered',color='orange')
plt.plot(T/365,0,'*',color='black')
plt.ylabel('Fraction of each category (normalized)',fontsize=9.5)
plt.xlabel('Time (years)',fontsize=9.5)
plt.legend(loc='upper right',fontsize=9.5)
plt.text(-2.4, 1.0, 'A', horizontalalignment='center', fontsize=15)
plt.show()
# plt.savefig('S and R.pdf') # (to save this figure, comment all other plots␣
↪and show() first)

# E & I:
plt.plot(TT,E,label='Exposed',color='blue')
plt.plot(TT,I,label='Infected',color='red')
plt.plot(T/365,0,'*',color='black')
plt.ylabel('Fraction of each category (normalized)',fontsize=9.5)
plt.xlabel('Time (years)',fontsize=9.5)
plt.legend(loc='upper right',fontsize=9.5)
plt.text(-2.8, 0.217, 'B', horizontalalignment='center', fontsize=15)
plt.show()
# plt.savefig('E and I.pdf') # (to save this figure, comment all other plots␣
↪and show() first)

# Minimum of I between peak1 and peak2:
y=int(365*1.4)
q=int(365*3)
# y and q: numbers to show two indeces: an approximation of the range of (peak1␣
↪and peak2)
# Let's plot this part:
plt.plot(TT[y:q],E[y:q],label='Exposed',color='blue')
plt.plot(TT[y:q],I[y:q],label='Infected',color='red')
plt.title("(Just to show the minimum of I in the range (peak1, peak2), it's not␣
↪important)")
plt.ylabel('Fraction of each category (normalized)',fontsize=9.5)
plt.xlabel('Time (years)',fontsize=9.5)
plt.legend(fontsize=9.5)
```

```python
print(f'Minimum of I between peak1 and peak2: {min(I[y:q])}')
```
